# Effectiveness of physical activity in obese children and adolescents and serum interleukin-10 levels: A protocol for systematic review

**DOI:** 10.1101/2024.02.29.24303561

**Authors:** Andreza Gomes Pascoal, Paula de Souza Mendes, Luiz Eduardo Rodrigues Lima, Bruno Mori

## Abstract

**Background:** Obesity is a global health problem that affects millions of people around the world, especially children and adolescents. The main causes of obesity are a sedentary lifestyle, unhealthy diet, and lack of reduction in the level of physical activity with negative consequences for physical and mental health, such as increased risk of cardiovascular diseases, diabetes, hypertension, dyslipidemia, and inflammation. Therefore, we will carry out a perform a systematic review to evaluate the effectiveness of physical activity in obese children and adolescents and serum interleukin-10 levels.

**Methods:** This protocol is based on recommendations from the Cochrane Collaboration on the steps of a systematic review. A literature search will be performed on the following database: PubMed, the Cochrane Library, LILACS (VHL), and Google Scholar. According to the Cochrane Risk of Bias Tool and the level of evidence for results, we will assess the quality of the included studies by using the Grading of Recommendations Assessment, Development, and Evaluation (GRADE) method. The Review Manager (v5.3) software will be applied to statistical analysis.

**Results:** From the study, we will evaluate the effectiveness of physical activity in obese children and adolescents and serum IL-10 levels correlated with age, ethnicity, sex, and body mass index (BMI).PROSPERO registration number: CRD42022355721

**Strengths and limitations of this study:** This article emphasizes the importance of evaluating the effectiveness of physical activity in obese children and adolescents and provides methodological guidance for evaluating clinical evidence. This study 1)provides a research protocol, 2) facilitates reasonable assessment of physical activity in obese children and adolescents and serum IL-10 levels 3) increases the potential importance of recommending physical activity in obese children and adolescents to improve serum levels of IL-10, which in turn has an immunoprotective effect. The lack of reports of these effects may be limited by the small sample size.

## Introduction

Regular physical activity has an inverse relationship with body weight and chronic non-communicable diseases, in addition to promoting benefits in physical fitness, it contributes to the maturation of the skeletal system, helps promote health, and improves the quality of life of children and adolescents, in addition to being essential for maintaining this habit in adulthood, therefore, an important determinant of the adolescent’s physical characteristics[1-2].

Several factors can influence physical activity patterns: individual characteristics, including motivation, motor skills, environmental characteristics, access to leisure spaces, architectural barriers, availability of time to practice physical activities, sociocultural support, and financial condition[3,4]. Another factor that may contribute to the increase in physical inactivity among young people is the adoption of sedentary behaviors such as watching TV and exposure to screens [5-9].

The current recommendation for physical activity in childhood and adolescence is 60 minutes or more of moderate physical activity daily, five or more days a week. However, intervention studies carried out in large urban centers demonstrate that more than 50% of children and adolescents do not meet current physical activity recommendations [10,11]. The reduction in energy expenditure (physical inactivity) in children and adolescents is the most important determinant of overweight and obesity, and it is not difficult to verify this variable in the lifestyle among children and adolescents. A systematic review and meta-analysis reveals robust evidence of a negative association between serum 25(OH)D levels <20 ng/ml and increased body mass index (obesity) and physical fitness levels [12].

Adipocyte-derived hormones play a fundamental role in regulating energy homeostasis, food intake, and many neuroendocrine functions. A state of leptin resistance has been suggested in obesity, characterizing the inflammatory process. Skeletal muscle was also recently identified as a secretory organ, the peptides produced, expressed, and released by muscle fibers were called myokines [2,12]. The factors that promote interference between adipose tissue and muscle cells and their impact on the regulation of energy homeostasis and metabolism are widely studied. However, it has been strongly suggested that physical inactivity promotes altered myokine responses, providing a potential mechanism for the association between obesity-associated sedentary behavior and many chronic diseases [13].

Interleukin 10 (IL-10) is an important anti-inflammatory and immunosuppressive cytokine that is produced by a variety of cells of the immune system, including T and B lymphocytes, monocytic cells, natural killer cells, dendritic cells, eosinophils, and neutrophils [14], is involved in several functions such as angiogenesis, lipolysis, control of glucose metabolism and regulation of inflammatory processes, demonstrating a protective effect against cardiovascular diseases and type 2 diabetes [15,16]. The chronic effect of physical exercise on inflammatory processes is a reflection of the sum of several anti-inflammatory actions promoted by the practice of physical exercise. Studies demonstrate that in response to physical effort, with the consequent triggering of an anti-inflammatory cascade, affected by the increase in circulating levels of IL-10 dependent on the practice of high-intensity physical activity, they can contribute to the chronic and subclinical inflammatory mediation caused by obesity. in children and adolescents. Therefore, we will carry out a systematic review and meta-analysis of RCTs and primary studies published to evaluate the effectiveness of physical activity in obese children and adolescents and serum interleukin-10 levels [17].

## 2. Materials and Methods

### 2.1 Study registration

The protocol for this systematic review has been registered in PROSPERO (CRD42022355721).

Besides, this protocol has been checked with preferred reporting items for systematic review and meta-analysis protocols (PRISMA-P) checklist [18].

### 2.2 Ethics

The approval of an ethics committee is not required for this protocol. Data are not individualized.

### 2.3 Inclusion criteria

2.3.1 Only clinical randomized controlled trials and primary articles to evaluate the effectiveness of physical activity in obese children and adolescents and the levels of serum interleukin-10, articles with children and adolescents who practice any type of physical activity that fits the parameters of obesity, articles that relate the practice of physical activity and interleukin-10 will be included.

2.3.2 Articles posted on PubMed, the Cochrane Library, LILACS (BVS), and Google Scholar.

2.3.3 Articles in all languages.

2.3.4 Articles published until October 2022.

2.3.5 Participants Children and adolescents, full-time schoolchildren, Children who agree to participate in the study and who have permission from their parents/guardians; Children who are in full physical ability, aged between 06 to 17 years, will be included. No restrictions on gender and race.

### 2.4 Exclusion Criteria

2.4.1 Children who do not have a consent form properly signed by their parents, as well as the Minor Consent Term, Children who have a physical or cognitive disability that does not allow them to perform the physical tests and who cannot answer the questionnaires.

2.4.2 Papers without complete data were not completed even after contacting the authors.

2.4.3 Secondary articles, such as systematic reviews with or without meta-analysis, overviews of systematic reviews.

2.4.4 Studies related to physical activity in obese children and adolescents but didn’t involve Interleukin-10.

### 2.5 Search strategy

A systematic search will be performed by PubMed, the Cochrane Library, LILACS (BVS), and Google Scholar. PubMed MeSH Terms: (Child) OR “Child” [Mesh] OR (Preschool) OR (Adolescents) OR “Adolescent”[Mesh] OR (Adolescence) OR (Teens) OR (Teen) OR (Teenagers) OR (Teenager) OR (Youth) OR (Youths) OR (Adolescents, Female) OR (Adolescent, Female) OR (Female Adolescent) OR (Female Adolescents) OR (Adolescents, Male) OR (Adolescent, Male) OR (Male Adolescent) OR (Male Adolescents) AND (Exercises) OR “Exercise”[Mesh] OR (Physical Activity) OR (Activities, Physical) OR (Activity, Physical) OR (Physical Activities) OR (Exercise, Physical) OR (Exercises, Physical) OR (Physical Exercise) OR (Physical Exercises) OR (Acute Exercise) OR (Acute Exercises) OR (Exercise, Acute) OR (Exercises, Acute) OR (Exercise, Isometric) OR (Exercises, Isometric) OR (Isometric Exercises) OR (Isometric Exercise) OR (Exercise, Aerobic) OR (Aerobic Exercise) OR (Aerobic Exercises) OR (Exercises, Aerobic) OR (Exercise Training) OR (Exercise Trainings) OR (Training, Exercise) OR (Trainings, Exercise) AND (obesity) OR “Obesity”[Mesh] OR (Obesity, Abdominal) OR (Obesity, Maternal) OR (Obesity, Metabolically Benign) OR (Obesity, Morbid) OR (Pediatric Obesity) AND (interleukin-10) OR “Interleukin-10”[Mesh] OR (Interleukin 10) OR (IL10) OR (IL-10) OR (CSIF-10) OR (Cytokine Synthesis Inhibitory Factor). Besides, ongoing trials will be retrieved from the Cochrane Library, LILACS (BVS), and Google Scholar. Details of the search strategy are stated as follows: (1) MeSH Terms:

Child; Preschool; Adolescents; Adolescence; Teens; Teen; Teenagers (2) Exercises; Physical Activity; Physical Activities; (3) obesity; (Obesity, Abdominal) (4) interleukin-10; Interleukin 10; IL10.Study titles and abstracts will be screened based on inclusion/exclusion criteria. We will cross-check the references cited as well. We will identify published studies until October 2022.

### 2.6 Interventions

We will include studies that use analysis of physical activity in any modality as the only intervention in the experimental group. Studies involving physical activity combined with other sports will be included if other activity practices are equally used in both experiments and control groups.

### 2.7 Outcomes

2.7.1 The primary result. The primary outcomes are mainly assessed by (1) levels of physical activity; (2) body mass index (BMI); (3) serum levels of interleukin 10; and (4) age.

2.7.2 The secondary outcome. Not applicable.

### 2.8 Data collection and analysis

#### 2.8.1 Study selection

All articles downloaded will be imported into Rayyan^®^ to remove the duplicated studies. According to inclusion and exclusion criteria, two reviewers will independently consult the title and abstract of every searched literature. Any disagreements on study selection will be resolved through discussion with other researchers. According to the Preferred Reporting Items for Systematic Reviews and Meta-Analyses (PRISMA) guidelines [18]. The flow diagram of all study selection procedures is shown in Figure 1. According to the Grading of Recommendations Assessment, Development, and Evaluation (GRADE) method, we will summarize the GRADE judgments [19].

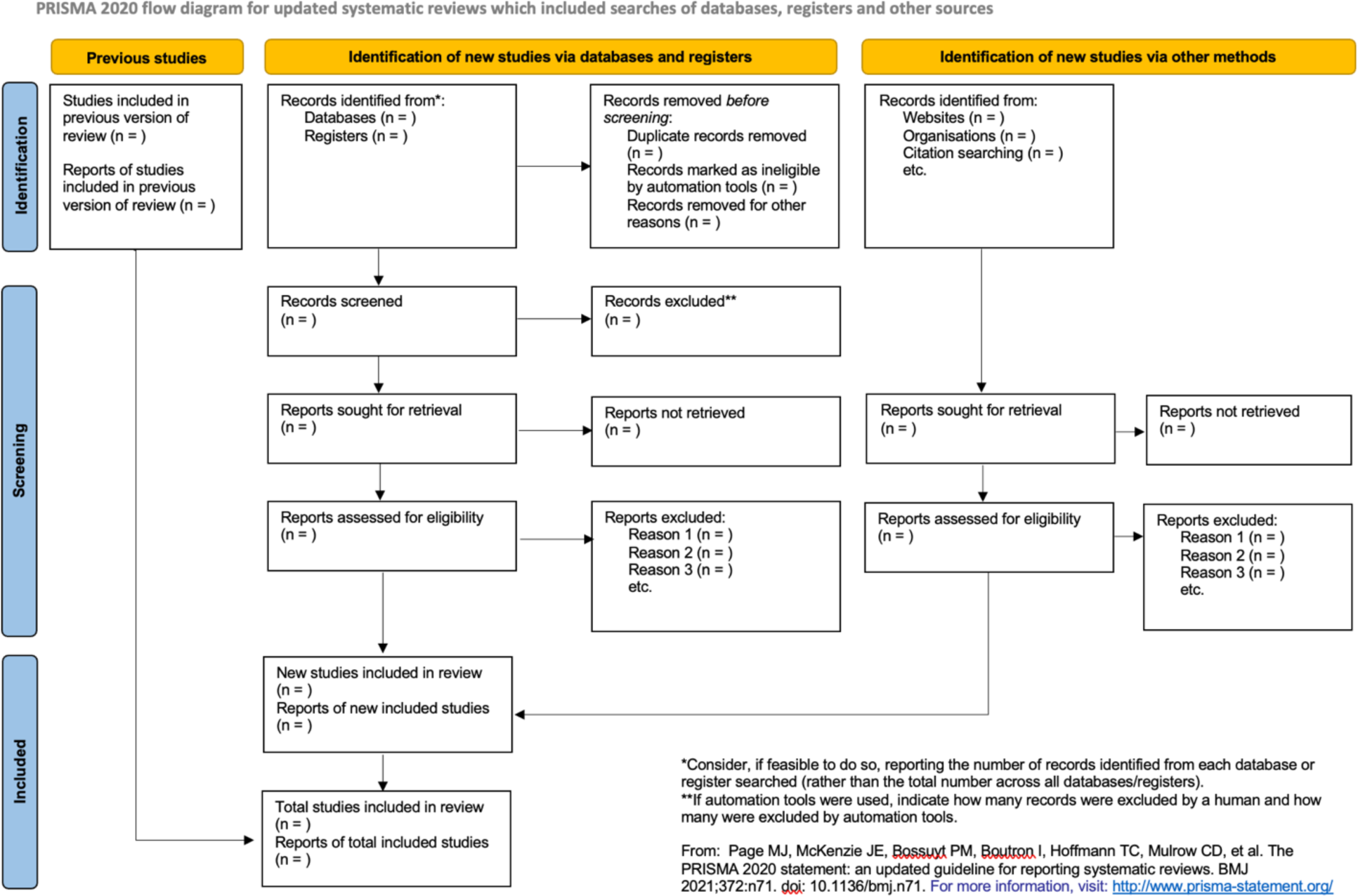

#### 2.8.2 Data extraction

Two independent authors performed the data extraction process and the following items from each trial were arranged. (1) The basic information of the trials or publications: the title of the article, first author, year, location, and language; (2) Participation baseline of the study. The subgroup condition and statistic distribution of the sample size, age, and sex included the study’s inclusion and exclusion criteria. Interventions in the observation group and the control group. Main outcome(s) and their scale basis. Besides, we will contact the lead author if there is any missing or unavailable information.

#### 2.8.3 Risk of bias assessment

We will assess the methodological quality of randomized controlled trials following the RCT quality assessment criteria recommended in the Cochrane Reviewer’s Handbook (5.3)[20]. Risk of The bias in Systematic Reviews (ROBIS) tool will evaluate the quality of reviews. Two assessors will check the results independently, and internal discussion will be performed if the results are inconsistent.

#### 2.8.4 Data synthesis

The data will be analyzed and synthesized through Review Manager (v5.3) software. Relative risk (RR) with 95% confidence intervals (CI) will be used for a dichotomous variable and weighted mean difference (WMD), or standard mean difference (SMD) with 95% CI for continuous variable. Cochrane Collaboration will be employed to compute the data synthesis.

#### 2.8.5 Assessment of Heterogeneity

I^2^ statistics will evaluate heterogeneity. If I^2^ ≥ 50% indicates the possibility of statistical heterogeneity, we will use a random-effects model for the calculation. Otherwise, a fixed-effects model will be used when the heterogeneity is not significant with its I^2^ < 50%.

#### 2.8.6 Subgroup analysis

When heterogeneity is high, subgroup analyses will be performed for different comparators separately if necessary. We will perform a subgroup analysis when heterogeneity is high. It can be discussed as gender type, physical activity duration, and IL-10 serum levels.

#### 2.8.7 Sensitivity analysis

The sensitivity analyses will be performed by excluding studies with high risks of bias and outliers that are numerically distant from the rest of the data.

#### 2.8.8 Publication bias

We will make funnel plots to evaluate the potential publication when studies included in the meta-analysis are more than ten. When an asymmetry of the funnel plot implies reporting bias, we will explain possible reasons.

### 2.9 Patient and Public Involvement

Patient and Public Involvement statements are not required since this is a meta-analysis-based on published studies.

## 3. Results

The results of this systematic review will be published as a peer-reviewed article.

## 4. Discussion

The practice of physical activity has been studied and correlated with a better quality of life in a broad sense, however, its anti-inflammatory effects have aroused common interest, given the metabolic modulation pathways [1,5]. Studies have observed an association between the level of physical activity and chronic non-communicable diseases (NCDs), obesity is a subclinical inflammatory aggravating factor [7]. Thus, detecting levels of physical activity among obese children and adolescents, as well as factors associated with physical inactivity, can contribute to the development of preventive programs that encourage the adoption of a physically active lifestyle with anti-inflammatory effects. [21,22].

Therefore, we will use a systematic review to evaluate the Effectiveness of physical activity in obese children and adolescents and serum interleukin-10 levels, to evaluate the general certainty of the evidence and the effects of physical exercise, potentially contributing to guidelines capable of implementing The results of this systematic review in clinical practice, the preferences of the monitoring team must also be considered, to determine if and when exercise should be the intervention of choice.

### Study Limitations

Currently, the conclusion of this systematic review may not be complete due to the relatively few studies that associate the practice of physical activity in obese children and adolescents and serum levels of IL-10 mechanism, the limited number of studies, as well as the limited number of participants included. Furthermore, the interpretation of statistical analysis can be difficult due to the limitations described. New studies with larger samples measuring obesity may be essential to obtain direct tips.

## 5. Conclusions

We hope that this study will provide relevant information about the effectiveness of physical activity in children and adolescents and serum levels of interleukin-10 and this conclusion will provide reliable evidence for more assertive clinical application. The conclusions of this review may further clarify the correlated immunobiological mechanism of physical activity and interleukin-10 expression to modulate the inflammatory effects of obesity in children and adolescents.

## Abbreviations

RCT: randomized controlled trials
GRADE: Grading of Recommendations Assessment Development and Evaluation
NCDs: Chronic Noncommunicable Diseases

## Author Contributions

Conceptualization, B.M and A.G.P.; methodology, B.M., and L.E.R.L.; software, B.M. and L.E.R.L.; validation, B.M.; formal analysis, B.M., L.E.R.L.; investigation, A.G.P.; resources, B.M.; data curation, B.M., A.G.P. and P.S.M.; writing—original draft preparation, B.M.; writing—review and editing, B.M.; supervision, B.M. All authors have read and agreed to the published version of the manuscript.

## Funding

No financing

## Institutional Review Board Statement

Not applicable. Informed Consent Statement: Not applicable.

## Data Availability Statement

The dataset generated during the current study is available from the corresponding authors upon reasonable request.

## Conflicts of Interest

The authors declare no conflict of interest.

## Notes

### Competing Interest Statement

The authors have declared no competing interest.

